# Functional and cognitive outcomes after COVID-19 delirium

**DOI:** 10.1101/2020.06.07.20115188

**Authors:** Benjamin C Mcloughlin, Amy Miles, Thomas Webb, Paul Knopp, Clodagh Eyres, Ambra Fabbri, Fiona Humphries, Daniel Davis

## Abstract

**Purpose:** To ascertain delirium prevalence and outcomes in COVID-19.

**Methods:** We conducted a point-prevalence study in a cohort of COVID-19 inpatients at University College Hospital. Delirium was defined by DSM-IV criteria. The primary outcome was all-cause mortality at 4 weeks; secondary outcomes were physical and cognitive function.

**Results:** In 71 patients, 31 (42%) had delirium, of which only 19 had been recognised by the clinical team. At 4 weeks, 20 (28%) had died, 26 (36%) were interviewed by telephone and 21 (30%) remained as inpatients. Physical function was substantially worse in people after delirium (−39 points on functional scale/166, 95% CI −92 to −21, p=0.01) (Table 2). Mean cognitive scores at follow-up were similar and delirium was not associated with mortality in this sample.

**Conclusions:** Our findings indicate that delirium is common, yet under-recognised. Delirium is associated with functional impairments in the medium-term.

**Key summary points:** 

**Aim:** To investigate functional and cognitive outcomes among patients with delirium in COVID-19.

**Findings:** Delirium in COVID-19 was prevalent (42%) but only a minority had been recognised by the clinical team. At 4-week follow-up, delirium was significantly associated with worse functional outcomes, independent of pre-morbid frailty. Cognitive outcomes were not appreciably worse.

**Message:** The presence of delirium is a significant factor in predicting worse functional outcomes in patients with COVID-19.

## Introduction

Delirium is one of the most common acute disorders in general hospitals, affecting around 25% of older patients [1]. Delirium is closely linked with adverse outcomes, including higher mortality, increased length of stay, long-term cognitive and functional decline, and risk of institutionalisation [2, 3]. Many screening instruments are available and the 4AT is the one best established within the UK National Health Service [4, 5]. Missed diagnoses may contribute to the excess mortality observed [6, 7], making systematic detection of delirium essential in any setting, no less so in the context of COVID-19 infection.

To date there has been limited work investigating the prevalence of and outcomes relating to delirium in COVID-19. Early studies describing the broad neurological features of COVID-19 suggest that 20–30% of hospitalised patients will present with or develop delirium or mental status changes, increasing to 60–70% in severe cases [8-10]. We set out to describe the point-prevalence of delirium in patients hospitalised with COVID-19, and quantify its association with mortality and cognitive and physical impairments at four weeks.

## Methods

### Study design and setting

We conducted a point prevalence study at University College Hospital of every inpatient (including critical care) with a diagnosis of COVID-19. All assessments for delirium took place on a single day, with outcomes measured four weeks later.

### Participants

We included all adult inpatients on Tuesday 21^st^ April 2020 who had tested positive for SARS-CoV-2 on combined throat and high nasal swab reverse-transcriptase polymerase chain reaction (RT-PCR). We did not include participants with a clinical suspicion of COVID-19 (e.g. on radiological or laboratory parameters) who were RT-PCR negative. We excluded patients who were discharged or died prior to the point of assessment.

### Outcomes

The primary outcome was all-cause mortality at four weeks. Deaths occurring outside of hospital were captured from the NHS Spine, a centralised national registry. Secondary outcomes were cognitive function and performance in activities of daily living at 4-week telephone follow-up. Cognitive function was measured using the modified Telephone Instrument for Cognitive Status (TICS-m), and performance in activities of daily living measured using a composite of the Barthel Index and Nottingham Extended Activities of Daily Living (NEADL) scores [11-13]

### Delirium assessments

All assessments were carried out on a single day by one rater (BM) with data reviewed by a delirium expert (DD). Delirium was defined by DSM-IV criteria. The 4AT was part of the assessment, with information supplemented by informant history (usually the clinical team) and review of medical notes from the previous 24 hours. Therefore, *disturbance of consciousness* was defined by altered arousal through use of the modified Richmond Agitation-Sedation Scale (mRASS) and/or inattention on ‘months of the year backwards’ or equivalent task; *change in cognition and/or perceptual disturbance* was identified through testing orientation using the AMT4 and components of the mental state examination; *fluctuating course* and *physiological basis* was determined by chart review. Where participants were unable to speak English (n=2), we made the diagnosis with the assistance of formal or family interpreters. All delirium cases were classified as hypoactive (reduced alertness), hyperactive (increased alertness or motor agitation), mixed (some features of both hypoactive and hyperactive), or no clear motor subtype.

### Other variables

We recorded additional clinical data: age, sex, ethnicity, dementia status (definite dementia = documented history; probable dementia = no documented diagnosis but history of progressive cognitive impairment affecting activities of daily living; no dementia). Clinical Frailty Scale (CFS) from 1 to 9 was determined by chart review by the geriatric medicine team. We recorded if patients had already been screened for delirium using any recognised tool and if a diagnosis of delirium had been recorded in the medical notes by the usual care team.

### Statistical methods

Differences between patients with and without delirium were analysed using χ^2^ tests for categorical data and independent t tests for continuous data. We defined cases of delirium as those only meeting all DSM-IV criteria; all other participants were considered to be ‘non-delirium’ unless they were completely unassessable because they were highly sedated in critical care. We compared 4-week survival in delirium versus non-delirium using logistic regression (primary outcome). For secondary outcomes, we treated TICS-m and Barthel+NEADL scores as continuous and compared these in people with and without delirium using linear regression, adjusted by age, sex and Clinical Frailty Scale score (as a continuous measure). Post-estimation procedures included examination of all residuals for heteroskedasticity. All analyses were conducted in Stata 14.0 (StataCorp, Texas).

## Results

A total of 82 patients were identified, though some were discharged prior to assessment (n=6), had died prior to assessment (n=3), or not present at review (n=2). The final sample included 71 patients. Among these, 25 patients were on acute medical wards, 5 in a High Dependency Unit, and 41 in critical care.

The mean age was 61 years (range 24 to 91), 51 (72%) were men, 6 (9%) had dementia or probable dementia and median clinical frailty score was 2 (IQR 2, 3) (Table 1). Delirium was identified in 31 patients (42%); hypoactive delirium accounted for 37% of presentations and 53% were hyperactive. Only a minority of cases (n=12, 39%) had been routinely recognised by the treating clinical team. Between delirium and non-delirium patients, 4AT sub-scores were different for each item (Supplementary Table). Where arousal was sufficient to assess directly, patients were evaluated for the presence of added symptoms. Phenomenologically, there were no differences in proportions with hallucinations, delusions, sleep disturbance or distress (Table 1).

**Table 1.** Patient characteristics of study participants and delirium status.

|  | No Delirium<br>(n=16) | Delirium<br>(n=31) | Unassessable<br>(n=24) | P |
| --- | --- | --- | --- | --- |
| Age (SD) | 63.3 (15.1) | 65.9 (15.3) | 55.5 (11.8) | 0.58 |
| Sex (%) M | 10 (19.6) | 25 (49) | 16 (31.4) | 0.33 |
| CFS (%) |  |  |  | 0.1 |
| 1 | 3 (18.8) | 5 (16.1) | 9 (37.5) |  |
| 2 | 9 (56.3) | 11 (35.5) | 12 (50) |  |
| 3 | 0 (0) | 5 (16.1) | 2 (8.3) |  |
| 4 | 1 (6.3) | 0 (0) | 0 (0) |  |
| 5 | 1 (6.3) | 1 (3.2) | 1 (4.2) |  |
| 6 | 1 (6.3) | 7 (22.6) | 0 (0) |  |
| 7 | 1 (6.3) | 2 (6.5) | 0 (0) |  |
| Dementia (%) | 1 (16.7) | 5 (83.3) | 0 (0) | 0.1 |
| Hallucinations (%) | 2 (50) | 2 (50) | 0 (0) | 0.07 |
| Delusions (%) | 0 (0) | 1 (100) | 0 (0) | 0.2 |
| Sleep disturbance (%) | 10 (62.5) | 6 (37.5) | 0 (0) | 0.68 |
| Distress (%) | 1 (33.3) | 2 (66.7) | 0 (0) | 0.33 |
| mRASS if no delirium |  |  |  | <0.01 |
| -5 |  | 1 | 6 |  |
| -4 |  | 0 | 7 |  |
| -3 |  | 2 | 6 |  |
| -2 |  | 1 | 2 |  |
| -1 |  | 5 | 1 |  |
| 0 |  | 3 | 1 |  |
| 1 |  | 3 | 0 |  |
| 4 |  | 1 | 0 |  |
| N/A |  | 15 | 1 |  |

Abbreviations: SD, standard deviation. *Unassessable* refers to patients with low mRASS. N/A refers to patients where mRASS was not required i.e ward patients.

At follow-up, 20 (28%) had died, 21 (30%) were still inpatients, 26 (36%) were interviewed by telephone and 4 (6%) could not be contacted (Figure 1). Of the remaining inpatients, 7 remained delirious and 8 were still unassessable.

**Figure 1.**
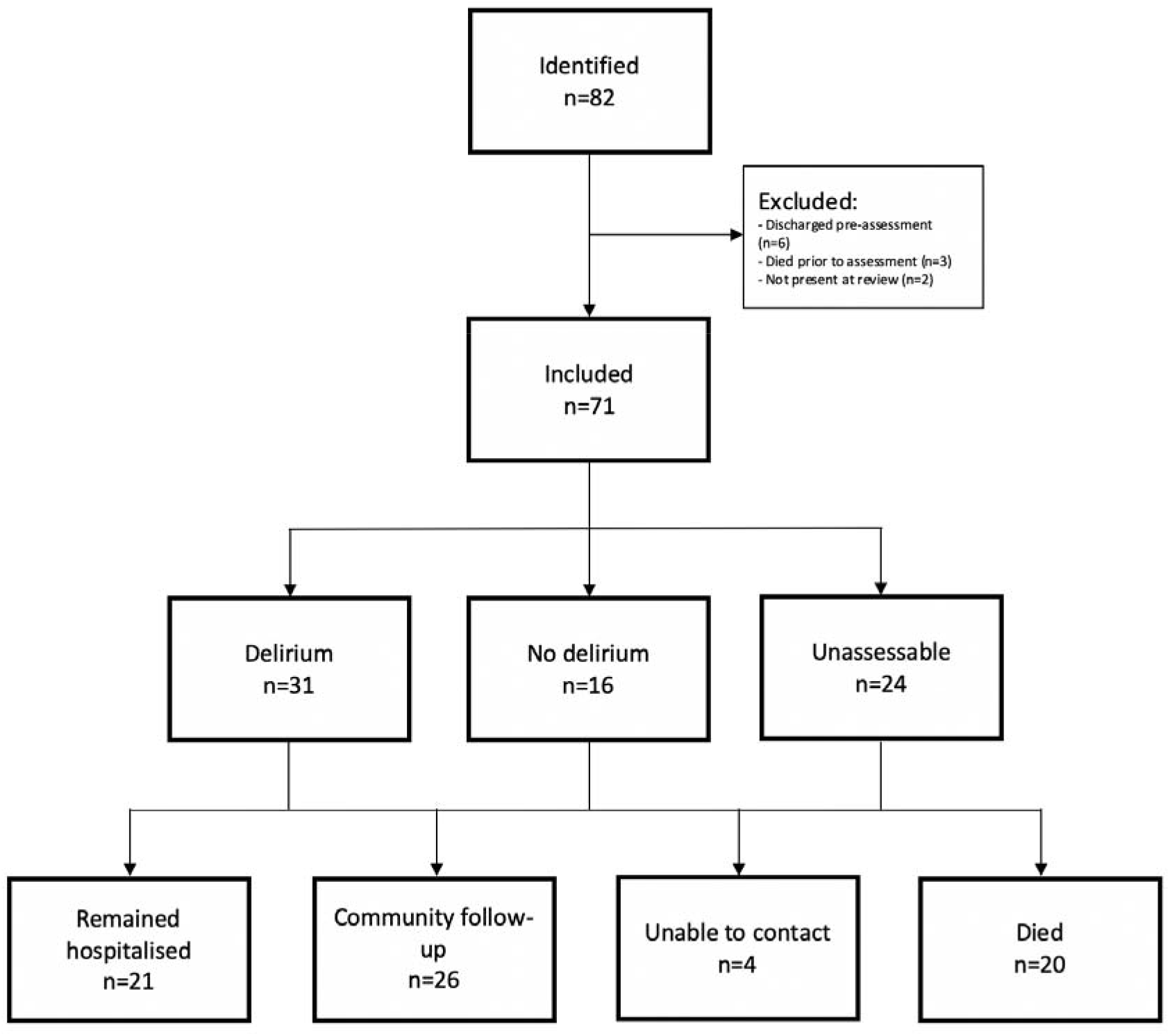
Flow chart describing patient recruitment, assessment and follow-up.

Mean cognitive scores at follow-up were similar in individuals with and without delirium (34.5 and 41.5, out of 53 respectively, p=0.06) (Figure 2). However, physical function was substantially worse in people after delirium (97 versus 153, p<0.01). These differences were still evident after adjustment for age, sex and pre-morbid frailty; here, delirium accounted for −39 out of 166 points (95% CI −92 to −21, p=0.01) (Table 2). Delirium was not associated with mortality in an age-sex-frailty adjusted model (OR 6.0, 95% CI 0.6 to 60, p=0.13).

**Figure 2.**
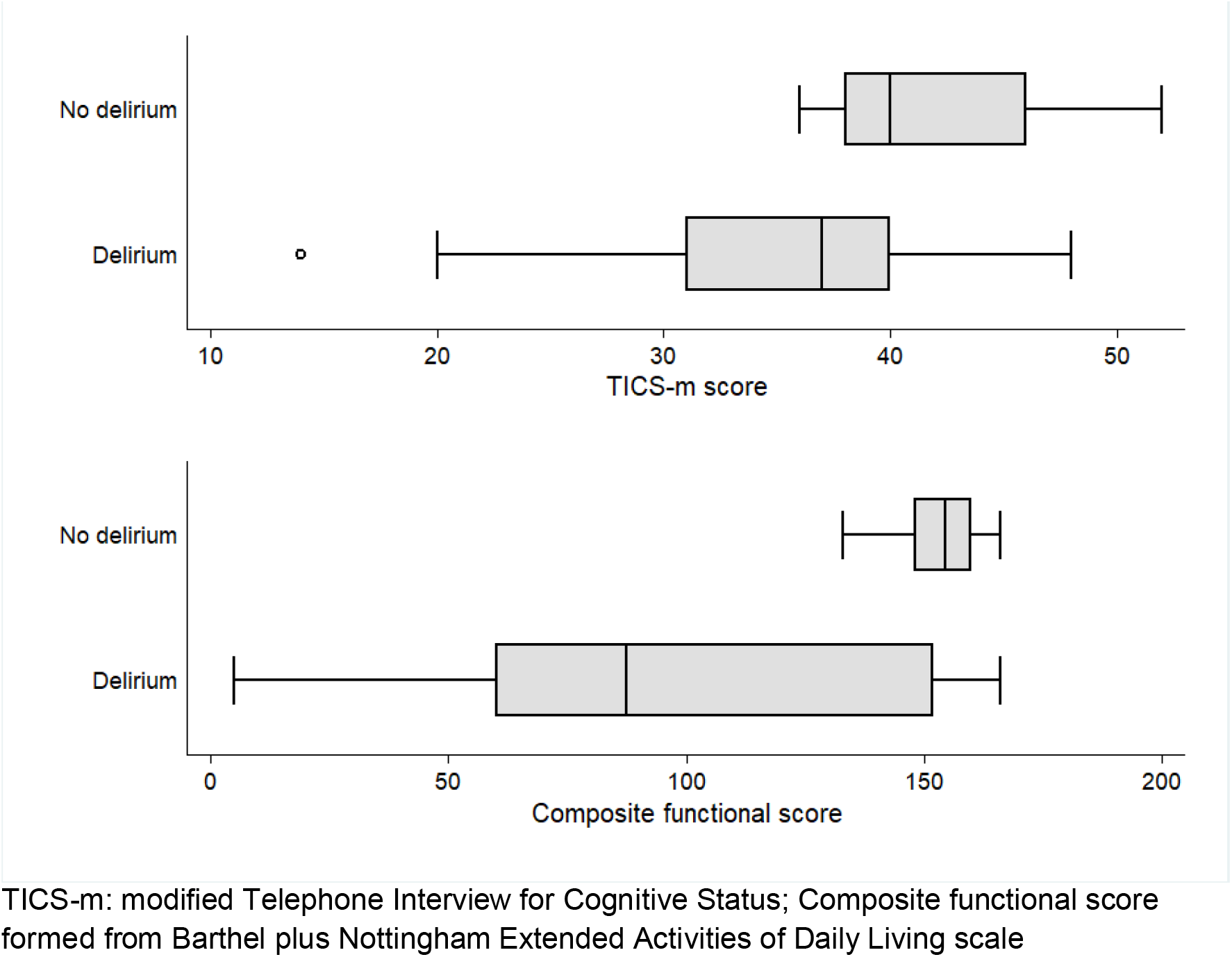
Cognitive and functional outcomes 4 weeks after delirium ascertainment

**Table 2.** Univariable and multivariable models estimating the associations with physical function at 4 weeks on a combined Barthel (100 points) and Nottingham Extended Activities of Daily Living score (66 points).

|  | Univariable models |  |  |  | Multivariable model |  |  |  |
| --- | --- | --- | --- | --- | --- | --- | --- | --- |
|  | HR | 95% CI |  | *P* | HR | 95% CI |  | *P* |
| Age | -1.3 | -2.6 | 0.08 | 0.06 | -0.6 | -1.6 | 0.5 | 0.25 |
| Sex | -40.2 | -92 | 11.5 | 0.12 | -16.2 | -51.6 | -19.2 | 0.34 |
| Delirium | -56.5 | -92.3 | -20.8 | 0.004 | -38.7 | -67.6 | -9.9 | 0.01 |
| CFS | -19 | -27.9 | -10.2 | 0.00 | -12.2 | -22 | -2.4 | 0.02 |

Abbreviations: HR, hazard ratio; CI, confidence interval; CFS, Clinical Frailty Scale

## Discussion

In patients hospitalised with COVID-19, delirium was found to be prevalent - but often undetected - and was associated with poor functional outcomes. We did not find many atypical features in this sample, though more hyperactive presentations were apparent than in other case series. There was no evidence of excess mortality or worse cognition at four weeks. Taken together, our findings suggest that delirium is a significant clinical complication of COVID-19 and long-term sequelae merits dedicated follow up.

Our results should be treated with caution. Data were collected at a single site in an urban university hospital and at a single time point, capturing a spectrum of stages in the disease course. As a sample of hospitalised patients, our findings may not be generalisable to community populations. With a substantial number of patients in critical care, there are additional complexities to the ascertainment of delirium. Our measure of physical function was established through self/informant report and direct assessments would have been more accurate. Nonetheless, our data are strengthened by the consistent and systematic approach to delirium detection and robust methods for follow-up.

These findings add to the growing body of work reporting the prevalence of and adverse outcomes associated with delirium (or ‘confusion’) in COVID-19 [8-10]. Delirium appears to be twice as common in COVID-19 than in other estimates (though these have often excluded patients in critical care) [14]. While adverse cognitive and functional outcomes from delirium are well-established, this is the first report to quantify these in the context of COVID-19. Though its impact seems to be clearer in terms of functional impairment, it is possible that persistent differences in cognitive outcomes would become more apparent with longer follow-up.

The pathophysiology of COVID-19 delirium, and its long-term outcomes, is likely to be multifactorial. Indirect mechanisms such as pyrexia, hypoxia, dehydration, metabolic derangements, and medications may be relevant. Direct pathways could also play a role, in the form of neuroinflammation and vascular injury [15]. These myriad risk factors underscore the importance of comprehensive assessment and management of delirium and its brain complications.

Our findings emphasise the requirement for dedicated delirium detection and management in COVID-19. Clearly, further work is needed to understand the mechanisms leading to delirium and its clinical and epidemiological outcomes. Though it is not known if any adverse sequelae could be mitigated through better delirium care, the scale and potential for distress itself justifies it as a clinical priority.

## Data Availability

On request

## Declarations

### Ethics approval

These analyses were conducted as part of a service evaluation project and individual consent was not necessary as determined by the NHS Health Research Authority (HRA), the regulatory body for medical research for England, UK. The HRA has the Research Ethics Service as one of its core functions and they determined the project was exempt from the need to obtain approval from an NHS Research Ethics Committee. https://www.hra.nhs.uk/about-us/committees-and-services/res-and-recs/

### Availability of data and material

on request

### Code availability

on request

### Authors’ contributions

BM, AM, TW, PK, FH, AF, CE collected the primary data. DD undertook the statistical analyses and had oversight of the project. BM drafted the first version of the manuscript. All authors contributed to revision and intellectual content of the final submission

## Acknowledgements

Thanks to Helen Bowden, MRC Unit for Lifelong Health and Ageing at UCL, who provided additional training.

---

**Supplementary Table.** 4AT in participants with and without delirium

|  |  | No Delirium (%) | Delirium (%) | P value |
| --- | --- | --- | --- | --- |
| Alertness |  |  |  | <0.01 |
|  | 0 | 16 (73%) | 6 (23%) |  |
|  | 4 | 0 (0%) | 8 (100%) |  |
| AMT4 |  |  |  | <0.01 |
|  | 0 | 14 (93%) | 1 (7%) |  |
|  | 1 | 1 (20%) | 4 (80%) |  |
|  | 2 | 1 (10%) | 9 (90%) |  |
| Attention |  |  |  | <0.01 |
|  | 0 | 14 (93%) | 1 (7%) |  |
|  | 1 | 2 (40%) | 3 (60%) |  |
|  | 2 | 0 (0%) | 10 (100%) |  |
| Acute change or fluctuating course |  |  |  | <0.01 |
|  | 0 | 16 (100%) | 0 (0%) |  |
|  | 4 | 0 (0%) | 14 (100%) |  |

